# Aggressive COVID-19 “second wave” in Italy

**DOI:** 10.1101/2020.11.11.20229872

**Authors:** Hua Zheng, Aldo Bonasera

## Abstract

A two-step model for the rise and decay of the COVID-19 is applied to the Fall resurgence of COVID-19 in Italy. Data starting from October 6, 2020 are compared to the same time interval in Italy starting from the complete lockdown on March 14, 2020. The model predicts more than 130,000 deceased by the end of the year 2020 if no effective measures are taken. If similar measures to the March ones are quickly adopted, the number of deceased may decrease to over 50,000. The situation is extremely serious and requires collaboration from everyone starting from wearing masks and other protections when social distancing is not feasible.

## 1 Introduction

In Refs. [1–3] we have introduced and applied a two-step model to the ‘first-wave’ of COVID-19 spread worldwide. After a decrease of cases in countries, which adopted effective measures to its spread [3], we observed a resurgence of COVID-19 in many western nations. In fact, differently from countries like China or S. Korea where the number of positives was effectively reduced to zero, in Europe a small number of cases was constantly registered everywhere. Such a small number, or probability (given by number of cases divided number of tests [1]) of about 10^−3^, acted as an infinitesimal seed to the exponential growth. In our model probabilities are described as [4–8]:

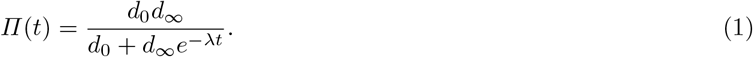

In the equation, *t* gives the time, in days, from the starting of the pandemic, or the time from the beginning of the tests for the virus. At time *t* = 0, *Π*(0) = *d*_0_ which is the very small value (or seed) from which the infection starts. In the opposite limit, *t* → ∞, *Π*(∞) = *d*_∞_, the final probability of affected people by the virus. If some effective measures are adopted (or a vaccine) then we may have a decrease of the probabilities given by:

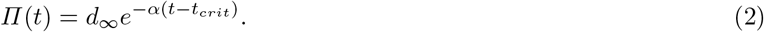

*α* and *t*_*crit*_ are fitting parameters. In order to derive the effective number of cases from the model we need to predict the number of tests performed daily for instance with power law fits:

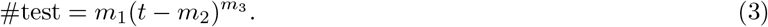

The parameters *m*_1−3_ are fitted to actual data in the period of interest. Multiplying eq. (2) or eq. (1) (depending on the stage of the spread, i.e., before or after a lockdown for instance) with eq. (3) gives the number of cases (positive or deceased) [3].

## 2 Results and prediction

In figure 1, we plot the number of cases as function of time starting from the specified days. The open symbols refer to the effective data recorded in Italy from March 14, 2020 (lockdown). After the rapid increase the data saturates and the total number of positives (deceased) can be estimated from the figure, 90 days after the lockdown. Further, we have collected the data with full symbols starting from October 6 up to November 20, i.e. at the time of writing. We have fitted the parameters entering eqs. (1) and (3) to the data (including the number of tests not shown here) from October 6 to the 29 (black symbols) and the predictions are given by the dashed lines. The blue points are actual data collected after October 29. A further fit to newer data results in an increase on the predictions [3]. A small increase of the data respect to the prediction maybe noticed and this is in part due to the increase of number of tests respect to our fit [3]. We notice that the new data increase even faster than in the Spring, which could be due to the number of tests performed in the two cases (we will discuss this in more detail in figure 2). The number of deceased displays the opposite behavior partially due to the fact that most probably COVID-19 deaths were easily identified, thus resulting on a weaker dependence on the number of tests. A large number of positives result in many deaths usually within 1-2 weeks time lag. We notice that the large increase of positives in the figure gives the exponential increase of the deceased, dashed lines. If no measures are taken the model predicts more than 1.7 millions positives and 130,000 deceased at the end of 2020.

**Fig. 1.**
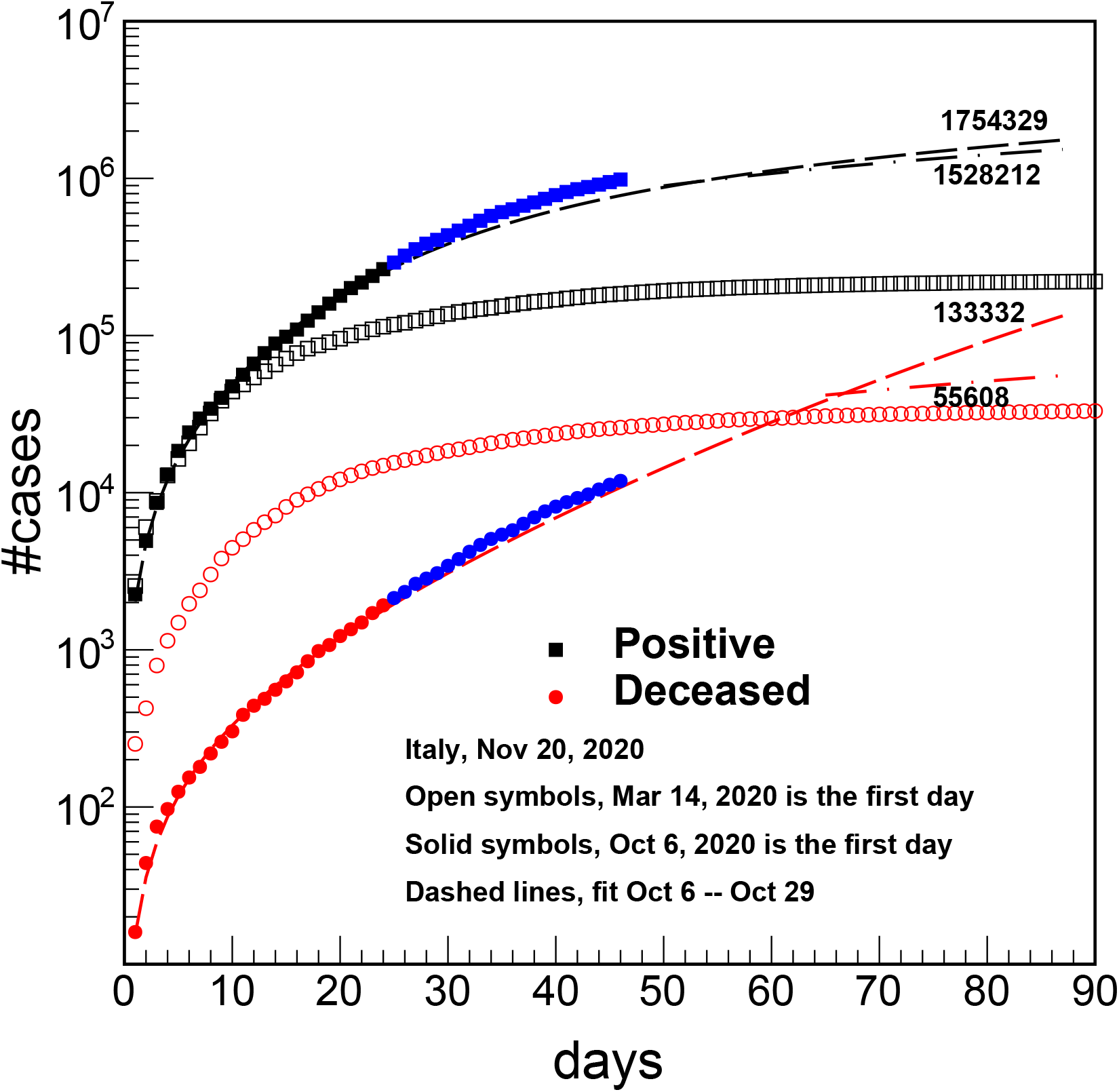
(Color online) Number of cases vs. time in days in Italy. The open symbols refer to the effective data registered starting from March 14^*th*^, 2020: the lockdown day. The full symbols refer to data starting from October 6^*th*^. The dashed lines are from the fits, eqs. (1-3).

**Fig. 2.**
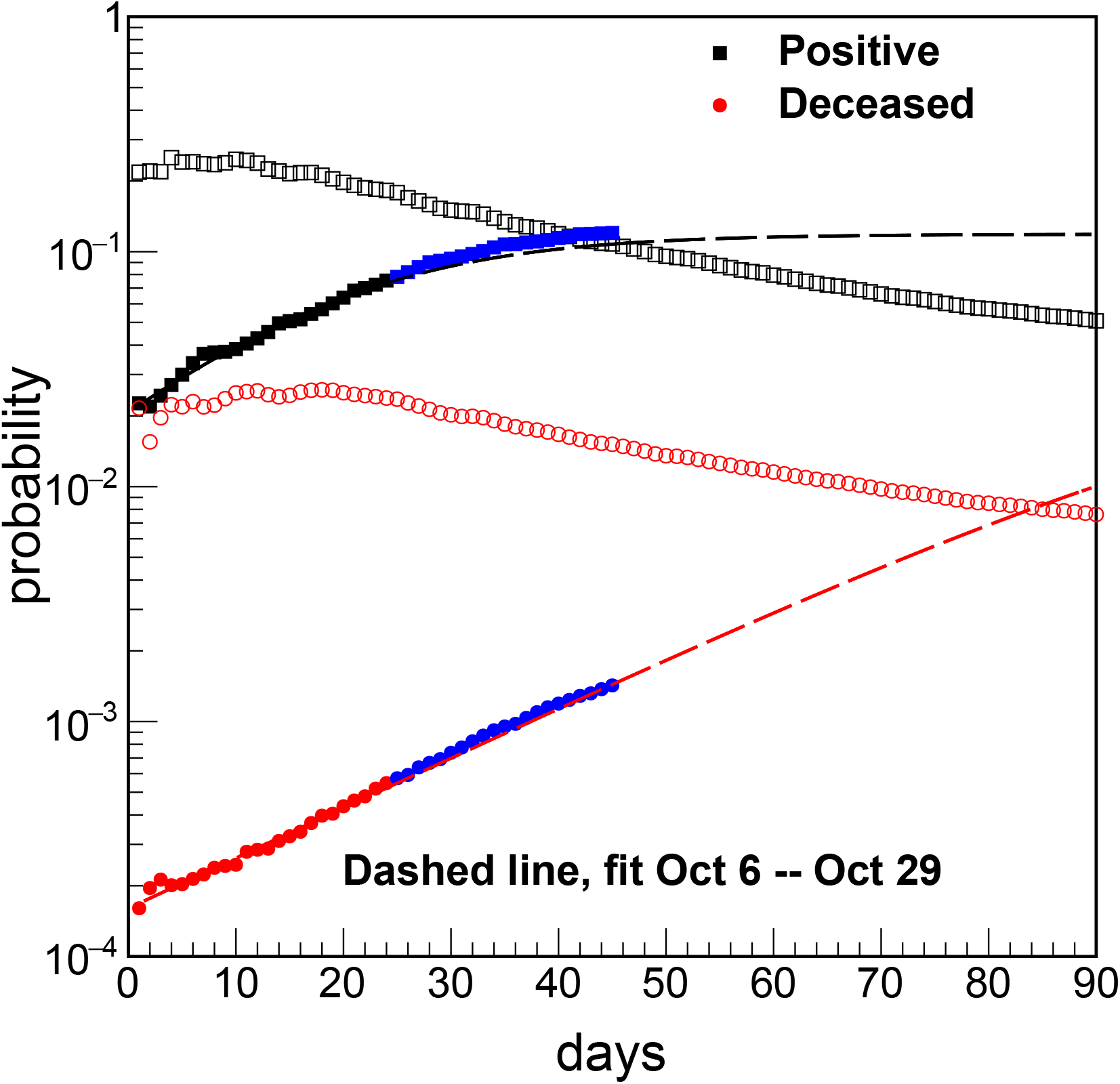
(Color online) Same as figure 1 but for the probabilities given by the number of cases divided the number of tests [1]. Fits are obtained using eq. (1), no decay stage is observed yet, opposite to the Spring data (open symbols).

Facing such a catastrophic situation, the Italian government will be forced to adopt new restrictive measures. At first the measures may be mild because the data does not show yet what is coming but eventually extremely severe measures will be adopted. If assuming that the measures will eventually have the same effects as in the spring of 2020 and use the same parameters for *t*_*crit*_=52.9 (66.7) and *α*=0.0198 (0.0168) for the positives (deceased), we will get a prediction which underestimates the data at the time of writing. Thus we choose *t*_*crit*_=52.9 (66.7) and *α* can be estimated from eq.(5) in Ref. [3]. Of course if there are further delays respect to our adopted *t*_*crit*_, or the measures are not as effective, the number of cases will go up. The predictions with dashed-dotted lines are displayed in the figure 1 and demonstrate a decrease in the number of cases by the end of the year 2020. Of course we cannot be sure that the exponential decrease will be the same as in the Spring because it refers to a different season and there may be a dependence on the weather etc. [9–12]. Also, we hope that the medical infrastructures have been strengthened, more personal protections, ventilators etc., are provided, which could modify the exponential decay especially for the deceased [2]. It is crucial that all people follow the suggestions spelled daily through social media to fight the virus but unfortunately refused by some.

Some of the features observed in figure 1 are due to the different number of daily tests. During the first wave the society was largely unprepared and the number of tests was not sufficient [1–3], thus the need to study probabilities. In figure 2 we plot the probabilities for the same cases of figure 1. Notice that, opposite to figure 1, probabilities are higher around March 14 due to the lower number of testing. Because of the lockdown, the exponential decrease described by eq. (2) occurred around March 24 [2, 3]. After 90 days the probabilities reached much lower values and continued to do so during the summer but, unfortunately, never reached zero. A similar behavior was observed for the deceased with a delay in the exponential decrease of about 7-10 days. The Fall data shows the exponential increase only and especially for the deceased there is no sign of flattening at later times. If no urgent measures are taken the situation will deteriorate quickly and in the figure 1 we have simulated a case if such measures are enforced around the middle of November 2020.

In order to better follow the spreading dynamics it is useful to plot the number of daily cases as in figure 3. These quantities can be obtained from actual data when available and from the time derivative of the two-step model. Notice that in the first ‘wave’ the number of daily cases reached their peaks approximately ten days after lockdown and later decreased exponentially. At the maximum, the number of deaths was about 1000/daily and could have reached much higher values without the adopted extreme measures. The current data is also showing an extremely rapid increase and it will continue to do so without lockdown. The theoretical prediction based on eqs. (1) and (3), i.e., without lockdown, predicts a power-law increase at later times as given by the fit to the number of tests. The final predictions with and without lockdown are displayed in figure 1. It is important to stress that without counter measures to the spread the number of positives may reach more than 20,000 daily, figure 3, which will seriously jeopardize health structures. If the adopted measures will be effective then the number of deceased should stabilize around 700 deceased daily. If not, we should observe a continue increase above a few thousands a day. We have plotted figure 3 in such a way that an interested reader or the authorities may update it daily to confirm or disprove the efficacy of the measures.

**Fig. 3.**
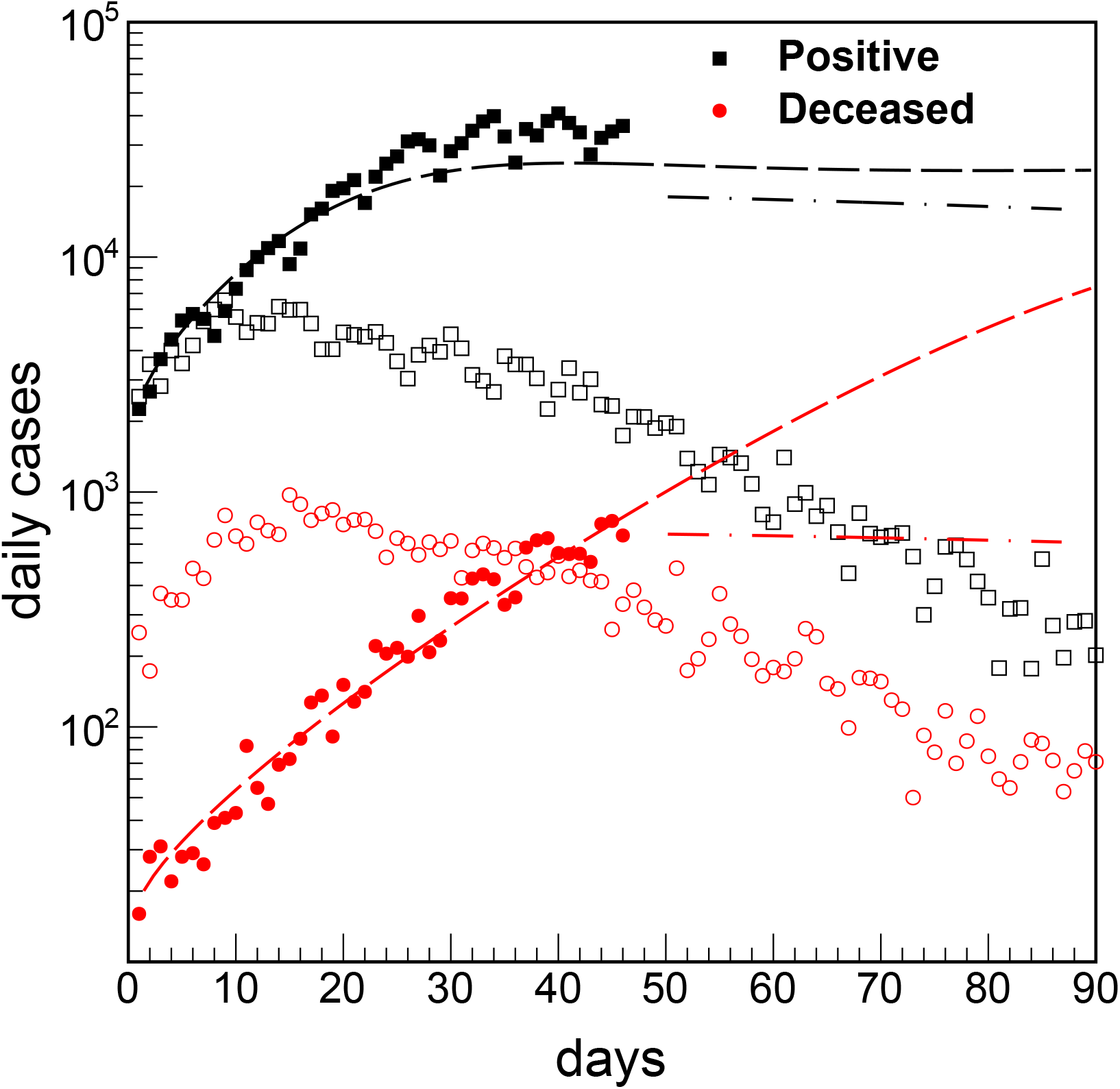
(Color online) Number of daily cases vs time, symbols as in figure 1. This quantity can be obtained from the time derivative of the quantities reported in figure 1 or from data directly (open and full symbols).

## 3 Conclusions

In conclusion, current data on the ‘second wave’ of COVID-19 and a two-step model depict a dramatic situation in Italy (and other European countries) which will result in tens of thousands deaths. The sooner the lockdown is adopted the more lives may be saved: this is no time to wait for miracles. One-week lockdown for the entire country will help to assess the situation in the different regions and, most importantly, contrast the large number of possible deaths, which we know from experience reaches its peak a few days after the peak in the number of positives. It is important to stress that without lockdown there is no peak but a rapid increase of cases as predicted in figure 1. Schools may go online if possible or just suspend lectures if not feasible. The lost period may be recovered in June or July if needed. After this preliminary lockdown, the situation maybe assessed in each region and measures can be taken accordingly. Safe ‘bubbles’ may be organized around factories, hospitals and other vital activities as discussed in Ref. [3]. Time is of essence, there is optimism that a vaccine may be ready soon but it will take sometime before its production and distribution. Thus it is imperative to slow down the spread of COVID-19 now with all means to save as many souls as possible.

## Supporting information

supplemental material for Italy+UK

## Data Availability

the data is available on request

## Acknowledgments

H. Zheng was supported by the Fundamental Research Funds for the Central Universities No. GK201903022.

