## supplemental material for Italy+UK for "Aggressive COVID-19 “second wave” in Italy"

### Covid-19 spread in Italy

**H.Zheng and A.Bonasera**

We monitor the second break of covid in Italy
Oct 6 is selected as the first day
The data are rescaled respect to Oct 6.

These are compared to data starting from March 14,2020 (lockdown)

**Last update: November 15,2020**

**The data sources are:**
**https://covid19.who.int**
**https://ourworldindata.org/grapher/full-list-total-tests-for-covid-19?time=2020-02-20..latest**

#### Relevant papers

https://doi.org/10.1101/2020.11.11.20229872

https://doi.org/10.1140/epjp/s13360-020-00811-z

https://doi.org/10.1140/epjp/s13360-020-00494-6

https://www.frontiersin.org/articles/10.3389/fphy.2020.00171/full

### Test number

**We fit two periods:**

1) **oct 5 — Oct 19, fitting parameters: 104529,4.77396e-13,1.07542**
2) **Oct 5 — Oct 25, fitting parameters: 95483.5,3.05395e-10,1.12018**

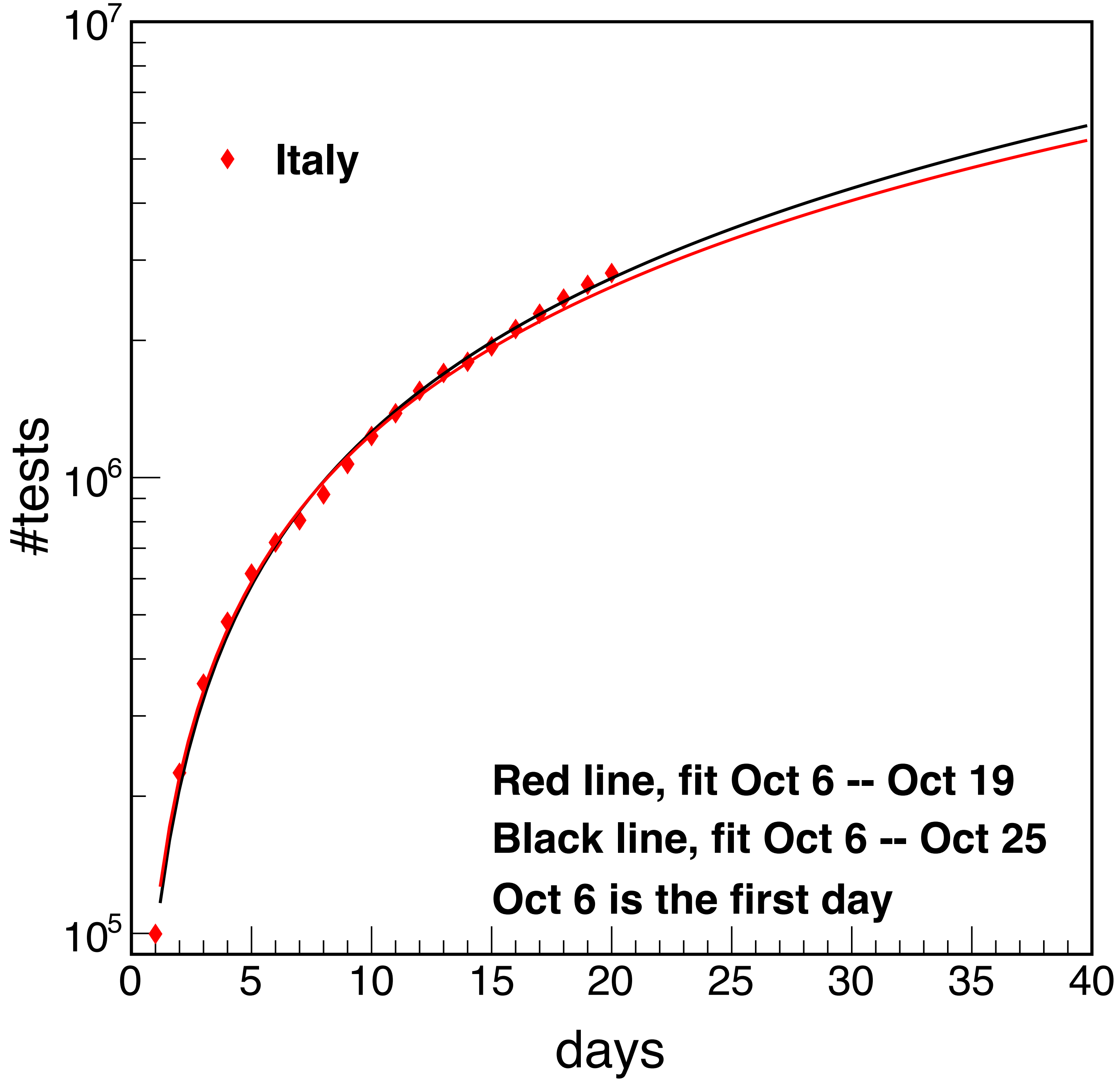


$$\#tests = m_1(t - m_2)^{m_3}$$

Check the probabilities

We fit two periods:

1) oct 5 — Oct 19, fitting parameters:
    1) pos 0.0285131,0.0589282,0.149018
    2) dec 0.000217757,0.000673943,0.0654084
2) Oct 5 — Oct 25, fitting parameters:
    1) pos 0.0263504,0.0848655,0.104863
    2) dec 0.000165612,0.011498,0.0479973

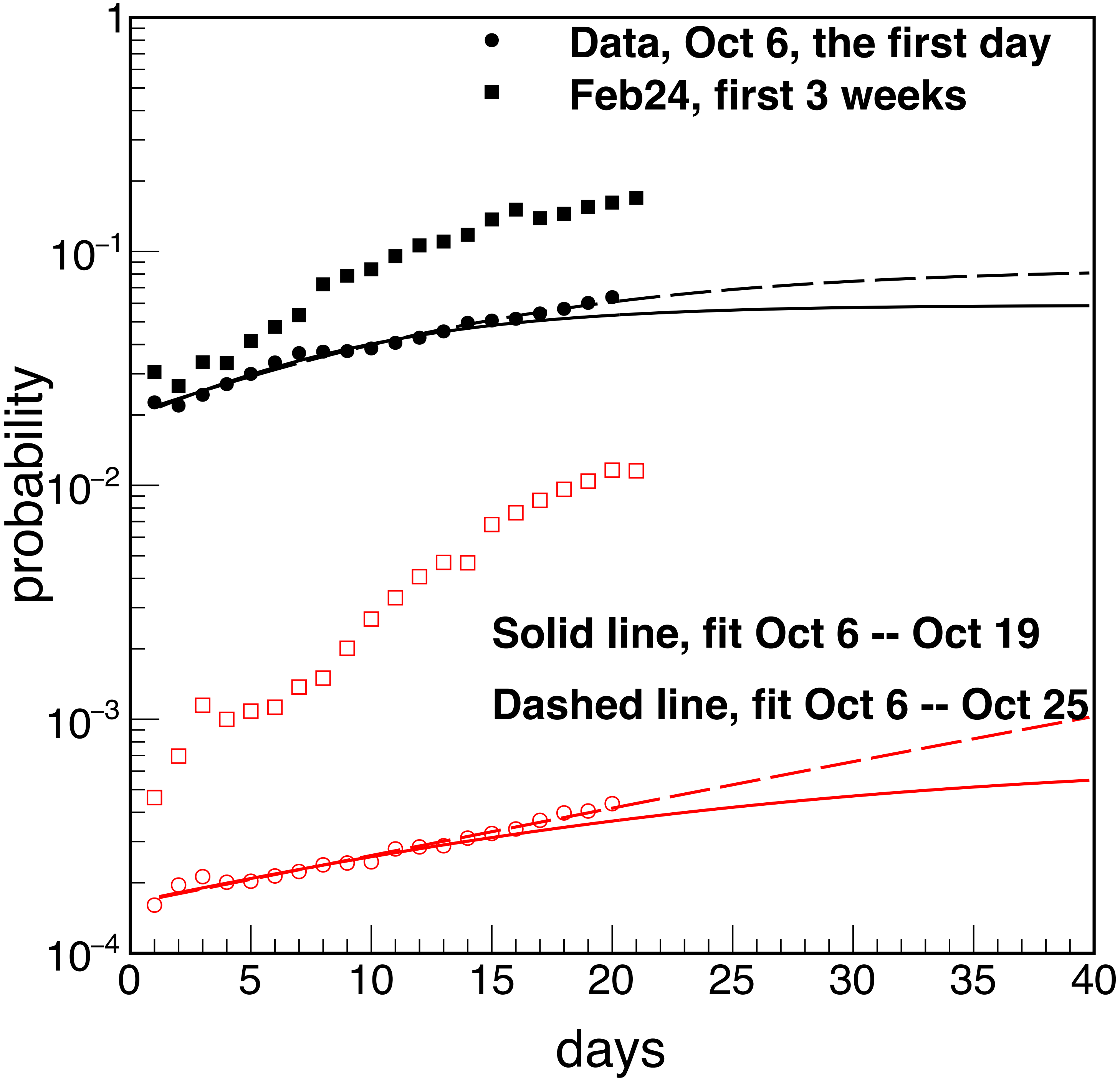


$$\Pi(t) = \frac{d_0 d_\infty}{d_0 + d_\infty e^{-\gamma t}}$$

### Prediction

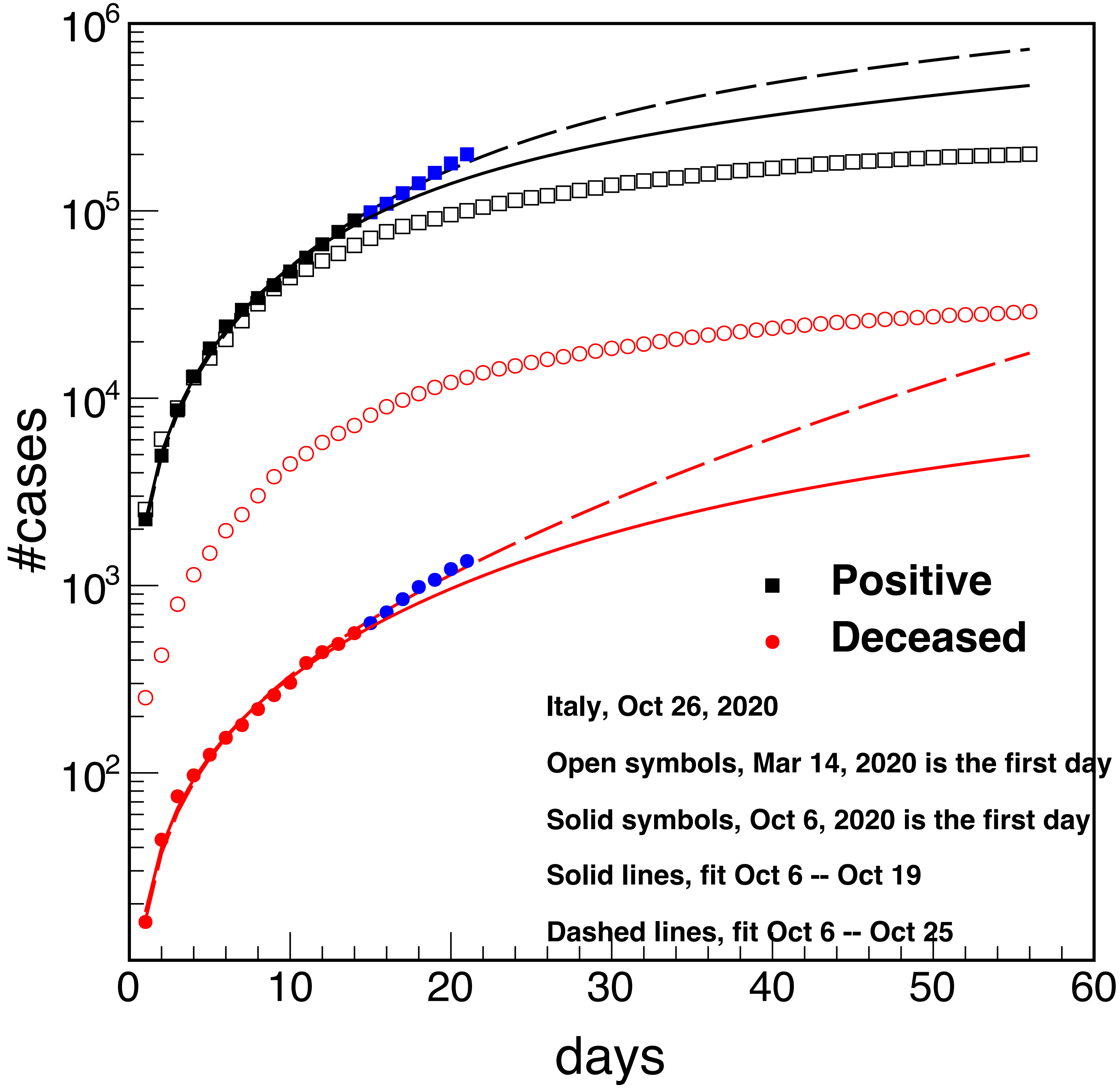


**Prediction for Nov 30, 2020**
**Fit oct 6 - oct 19**
**Pos 467071**
**Dec 4951**

**Fit oct 6 - oct 25**
**Pos 729496**
**Dec 17426**

$$\#cases = \frac{d_0 d_\infty}{d_0 + d_\infty e^{-\gamma t}} m_1 (t - m_2)^{m_3}$$

### Extend the prediction to Dec 31st, 2020

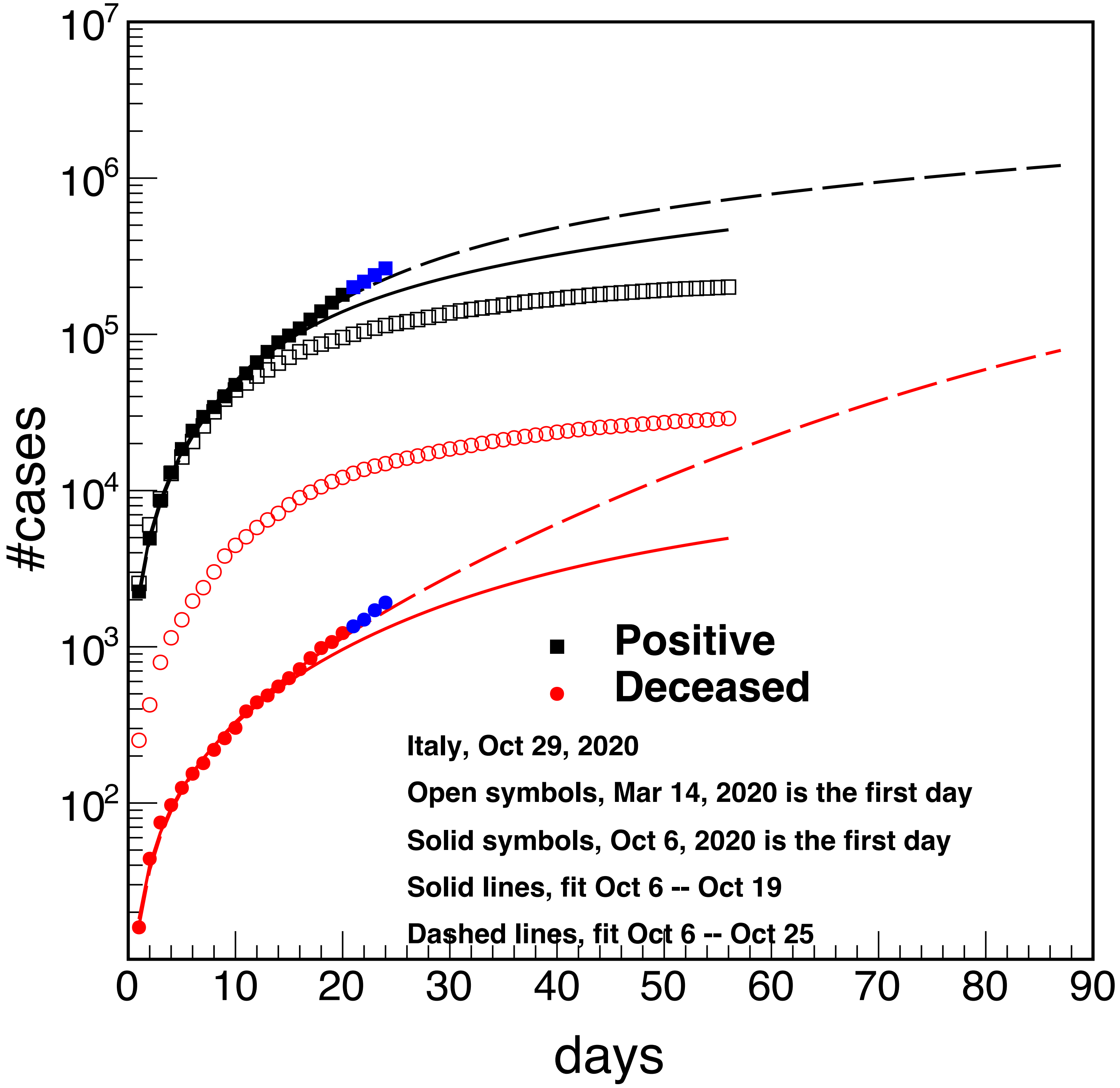


**Dec 31st**

**Pos 1205362**

**Dec 79049**

$$\#cases = \frac{d_0 d_\infty}{d_0 + d_\infty e^{-\gamma t}} m_1 (t - m_2)^{m_3}$$

**The blue (black&red-used for the fit) points are the real data which can be compared to the prediction. Data are a little higher than our prediction, but the trend is the same. We need to see how the real data evolve in the coming days.**

**Include the second stage using the same parameters of reference [1]. This assumes complete lockdown by the middle of November, similar to March 14, 2020.**

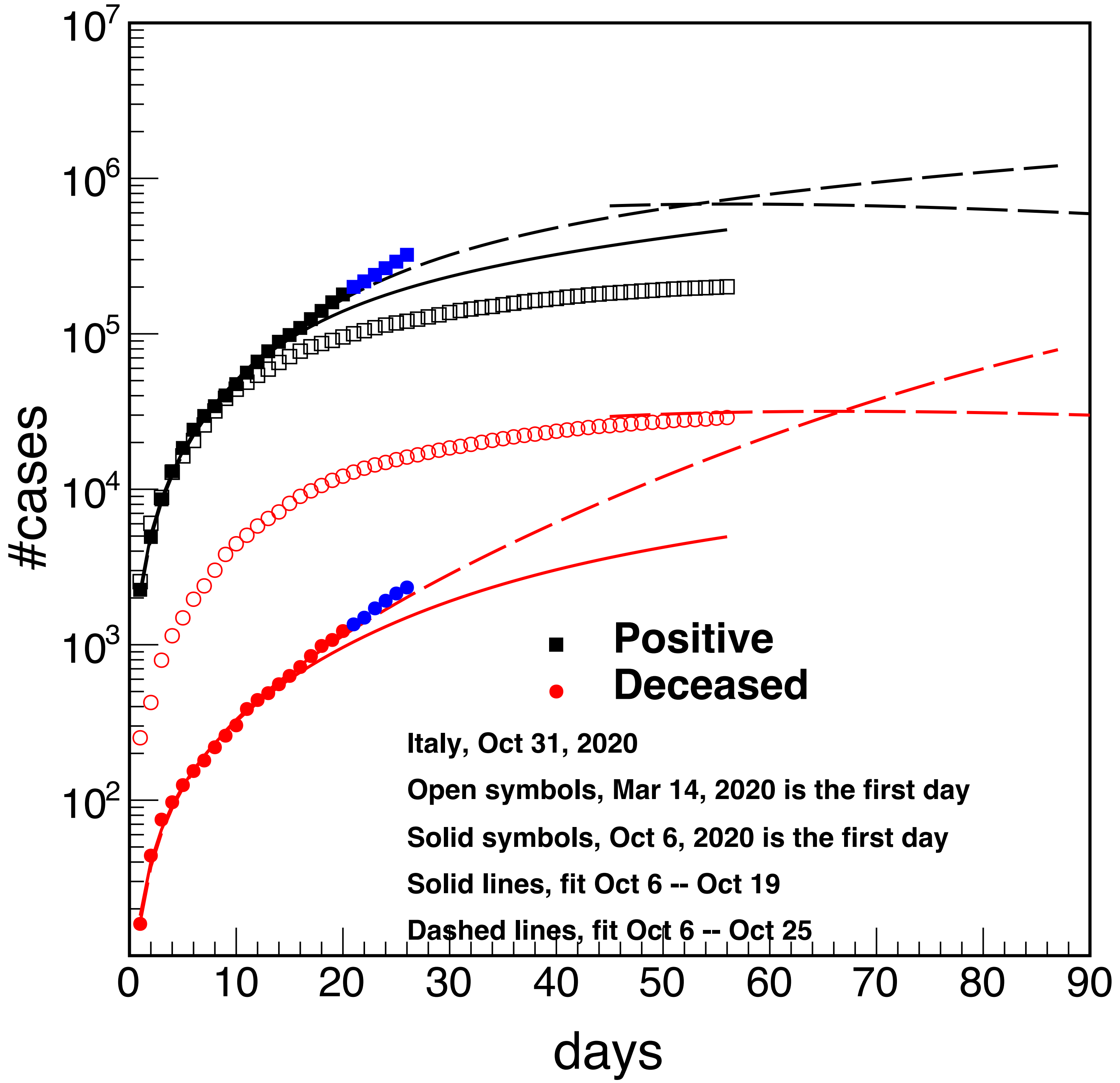


**The deceased is not convergent yet, needs more data**

$$\#cases = \frac{d_0 d_\infty}{d_0 + d_\infty e^{-\gamma t}} m_1 (t - m_2)^{m_3}$$

**Extend the prediction to Dec 31st, 2020. Adjust the number of tests since they are increasing respect to our previous fit.**

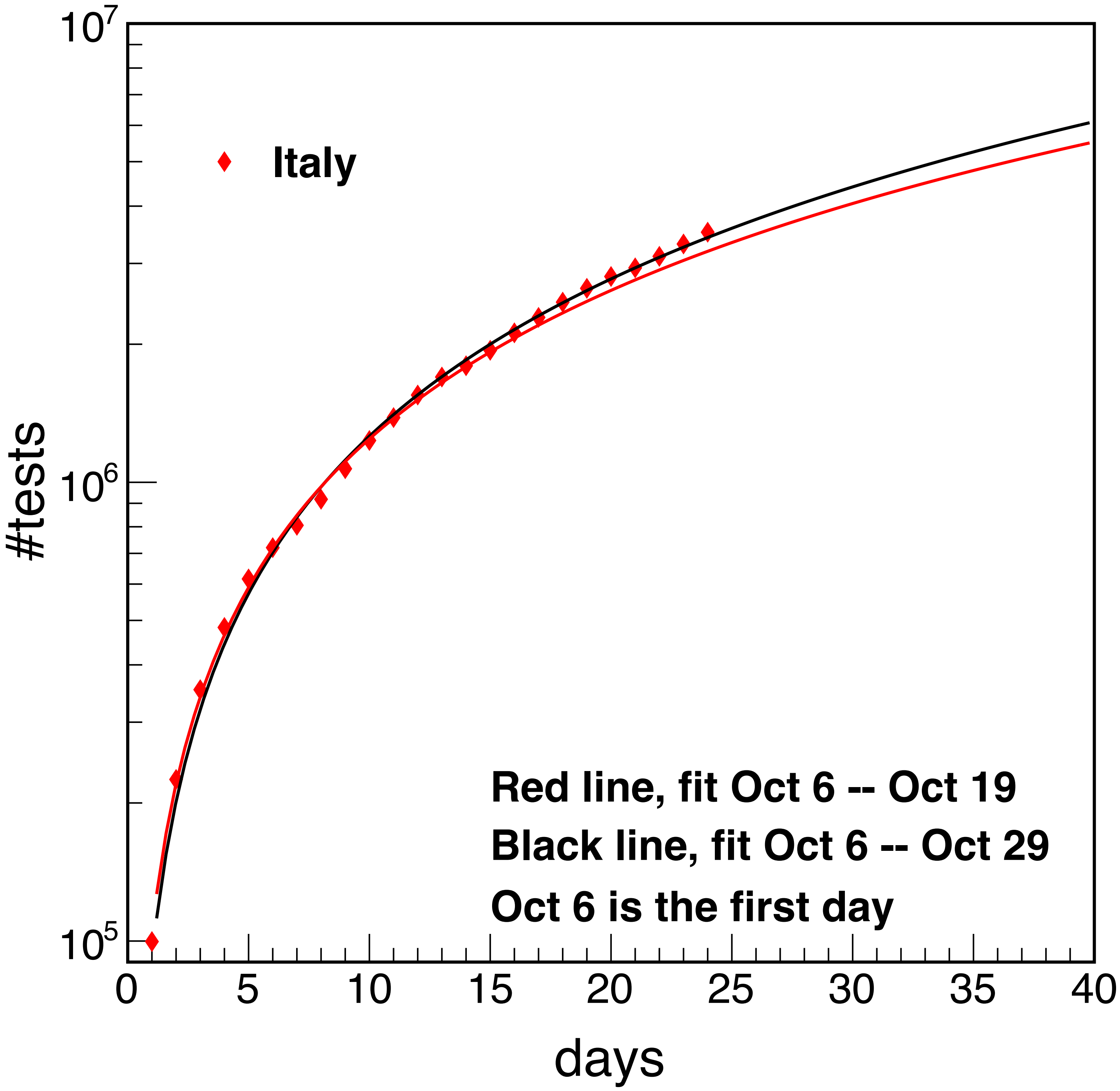


**Fit the period oct6-oct29**

**91472.1,9.94205e-12,1.13916**

$$\#tests = m_1(t - m_2)^{m_3}$$

### Extend the prediction to Dec 31st, 2020

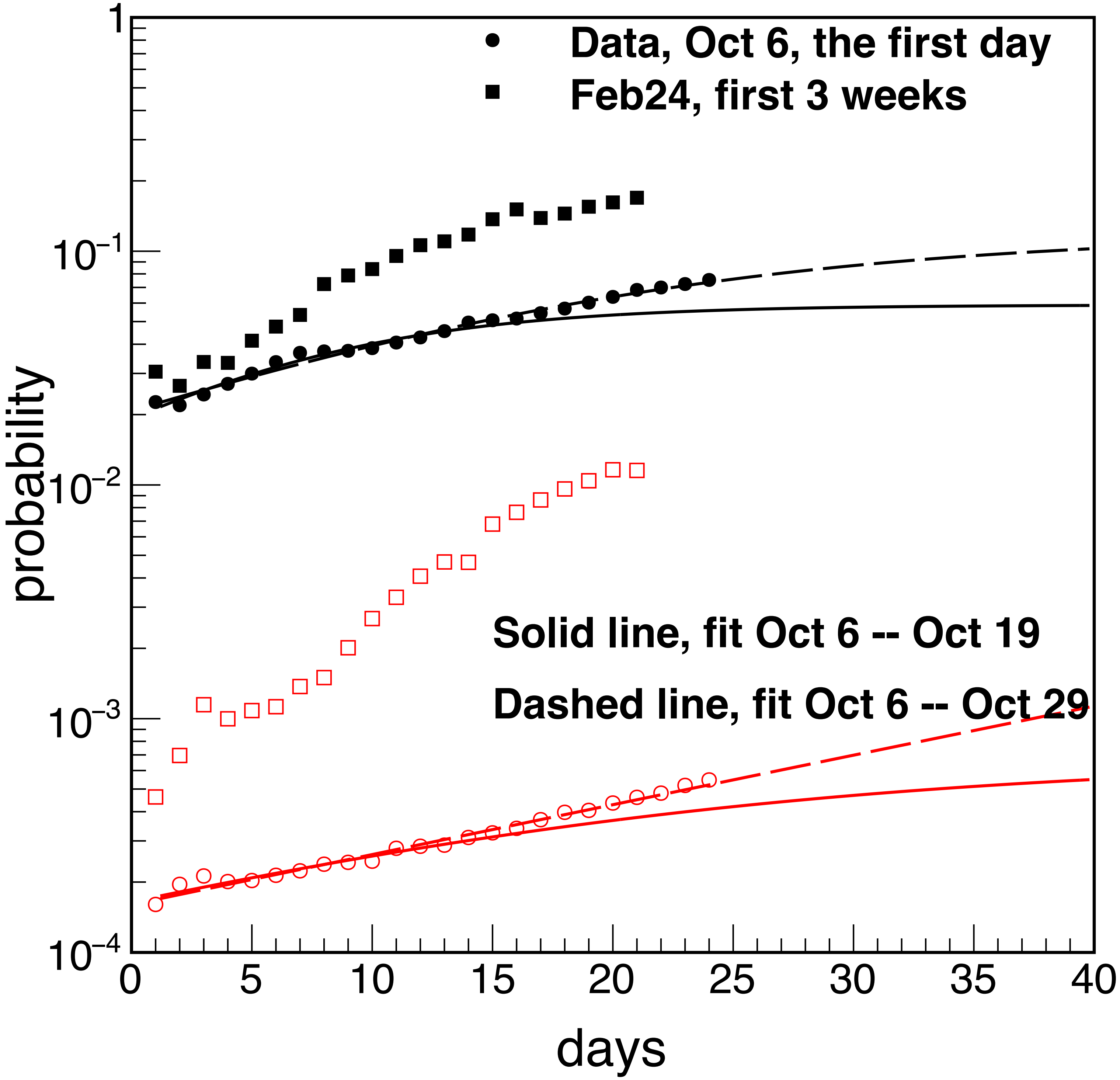


**Probability fit**

**Fit the period oct6-oct29**

$$\Pi(t) = \frac{d_0 d_\infty}{d_0 + d_\infty e^{-\gamma t}}$$

### Extend the prediction to Dec 31st, 2020

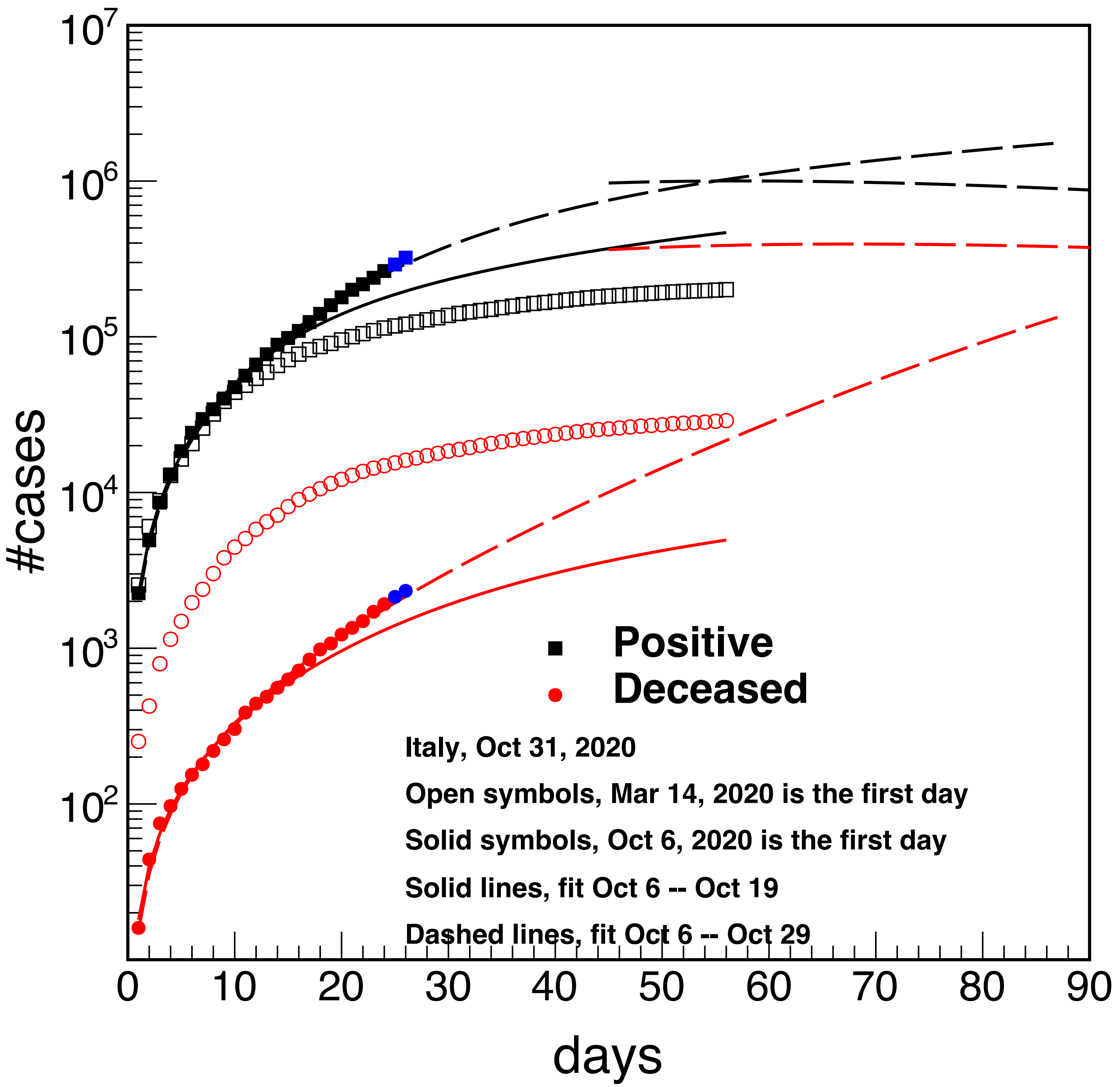


**Dec 31st**

**Pos 1754329**

**Dec 133332**

$$\#cases = \frac{d_0 d_\infty}{d_0 + d_\infty e^{-\gamma t}} m_1 (t - m_2)^{m_3}$$

### Extend the prediction to Dec 31st, 2020

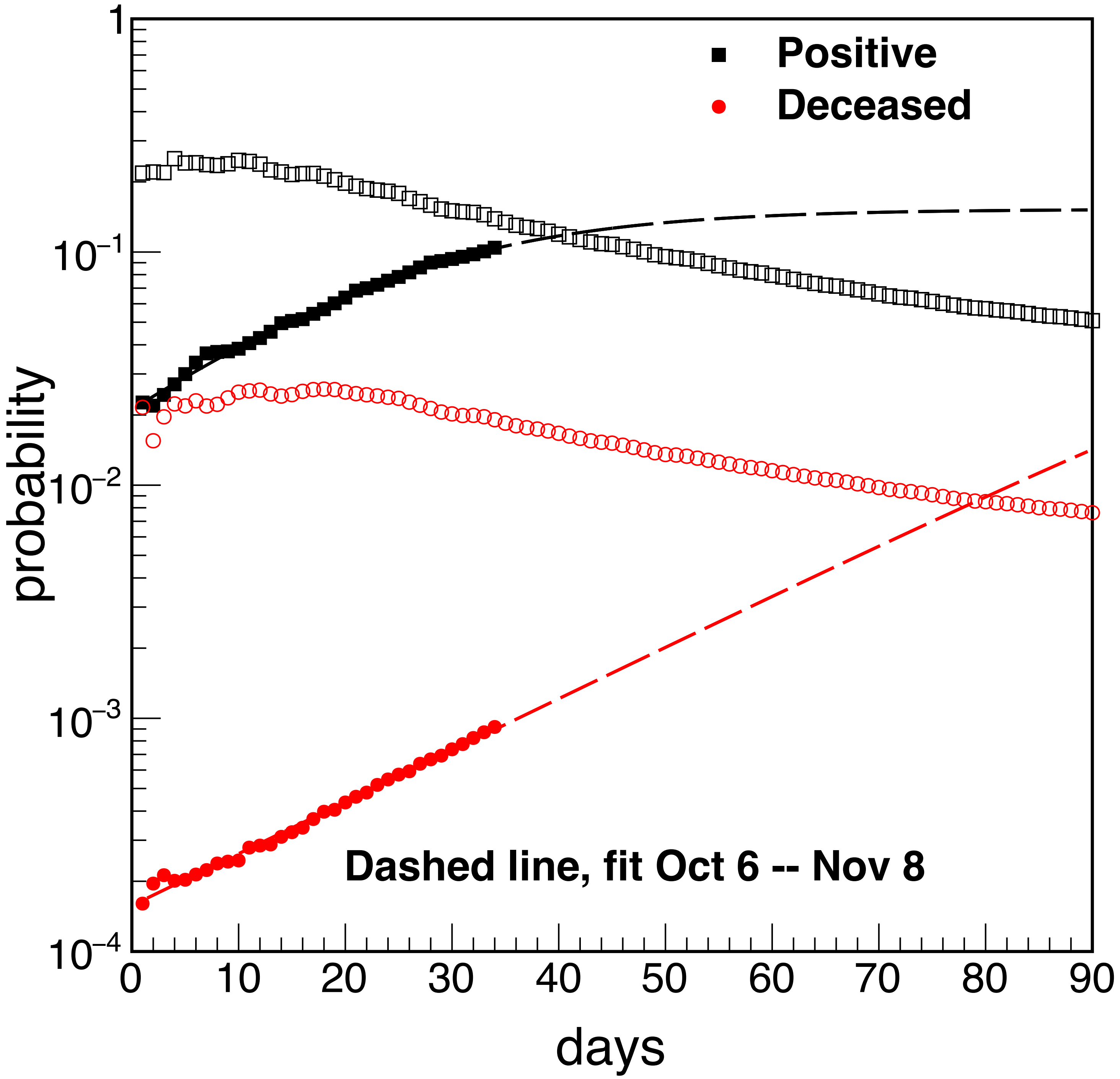


**Probability fit**
**Fit the period oct6-nov8**
**Pos 0.0245332,0.152631,0.0756671**
**Dec 0.000157569,0.136735,0.05126**

**Test number fit**
**79634.2,0,1.19448**

$$\Pi(t) = \frac{d_0 d_\infty}{d_0 + d_\infty e^{-\gamma t}}$$

**open symbols refer to data starting from March 14, 2020 (lockdown)**

**New prediction for Dec 31st (fit Oct 6 - Nov 8)**

**Positives 2498830 (no lock down) 1261352(lockdown)**

**Deceased 204564 (no lock down) 56107(lockdown)**

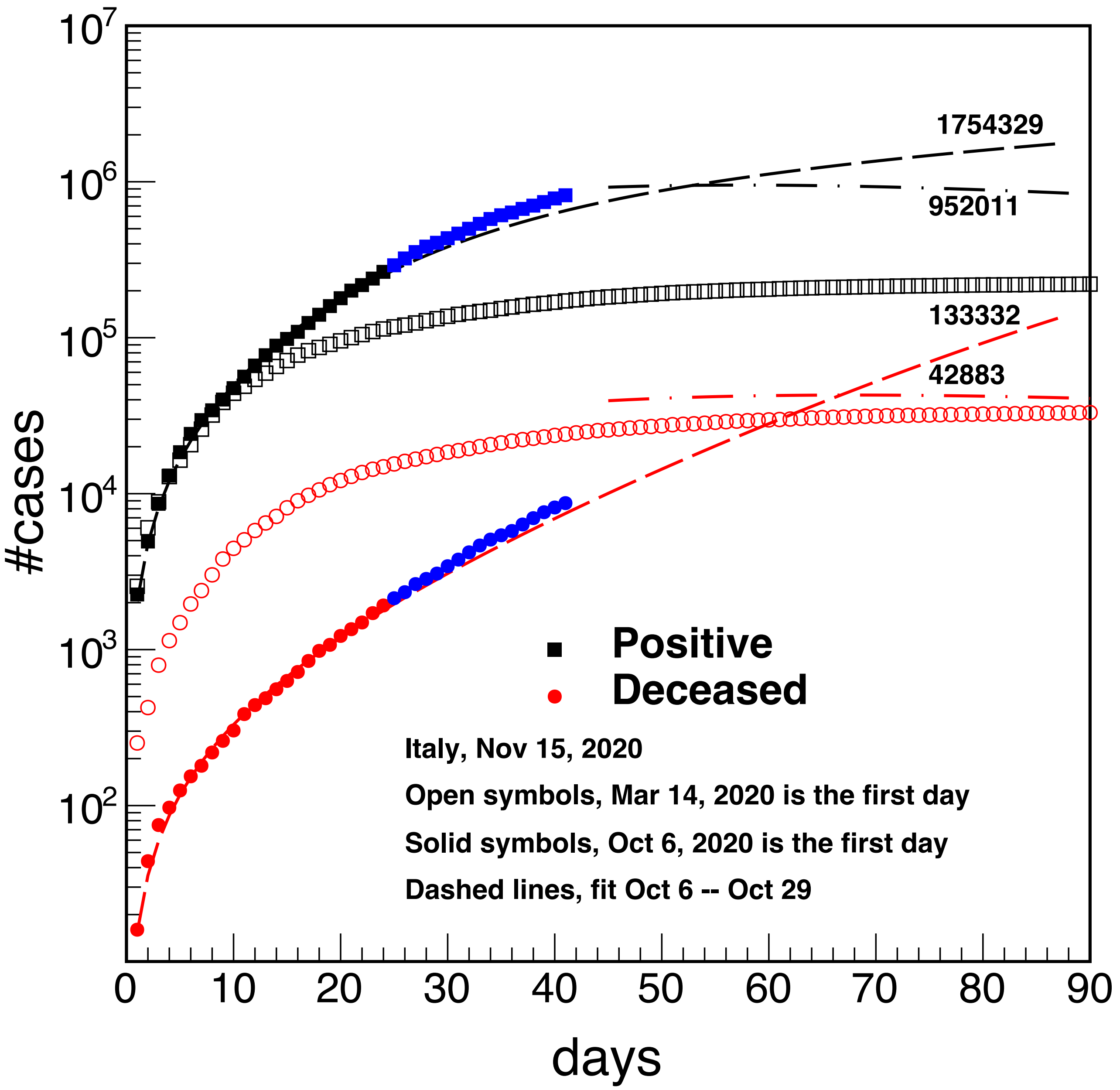


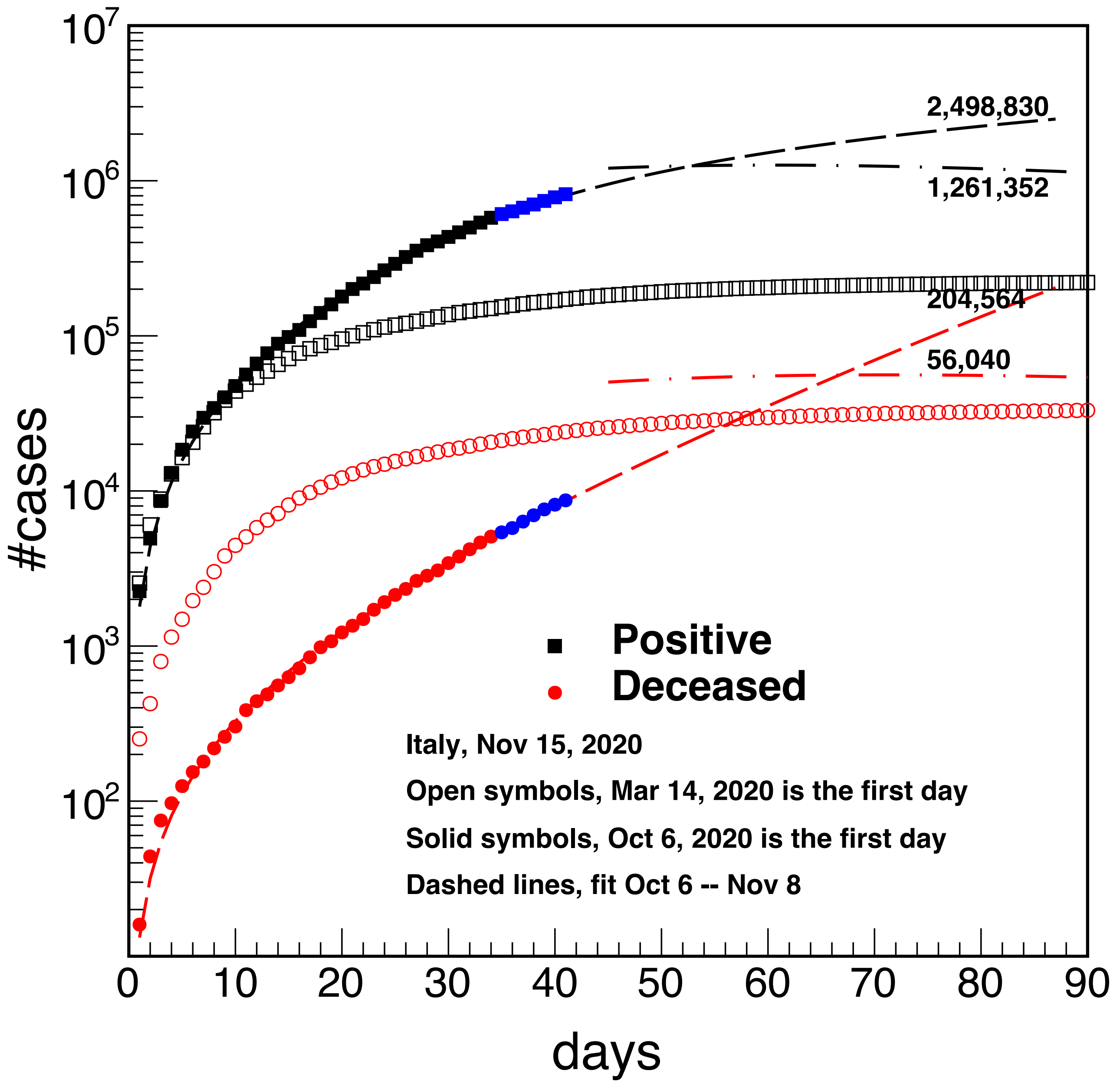


**Two fitting period 1) oct6-oct29 (24 days), 2) oct6-nov8 (34 days)**

$$\#cases = \frac{d_0 d_\infty}{d_0 + d_\infty e^{-\gamma t}} m_1 (t - m_2)^{m_3}$$

#### Two fitting period 1) oct6-oct29 (24 days), 2) oct6-nov8 (34 days)

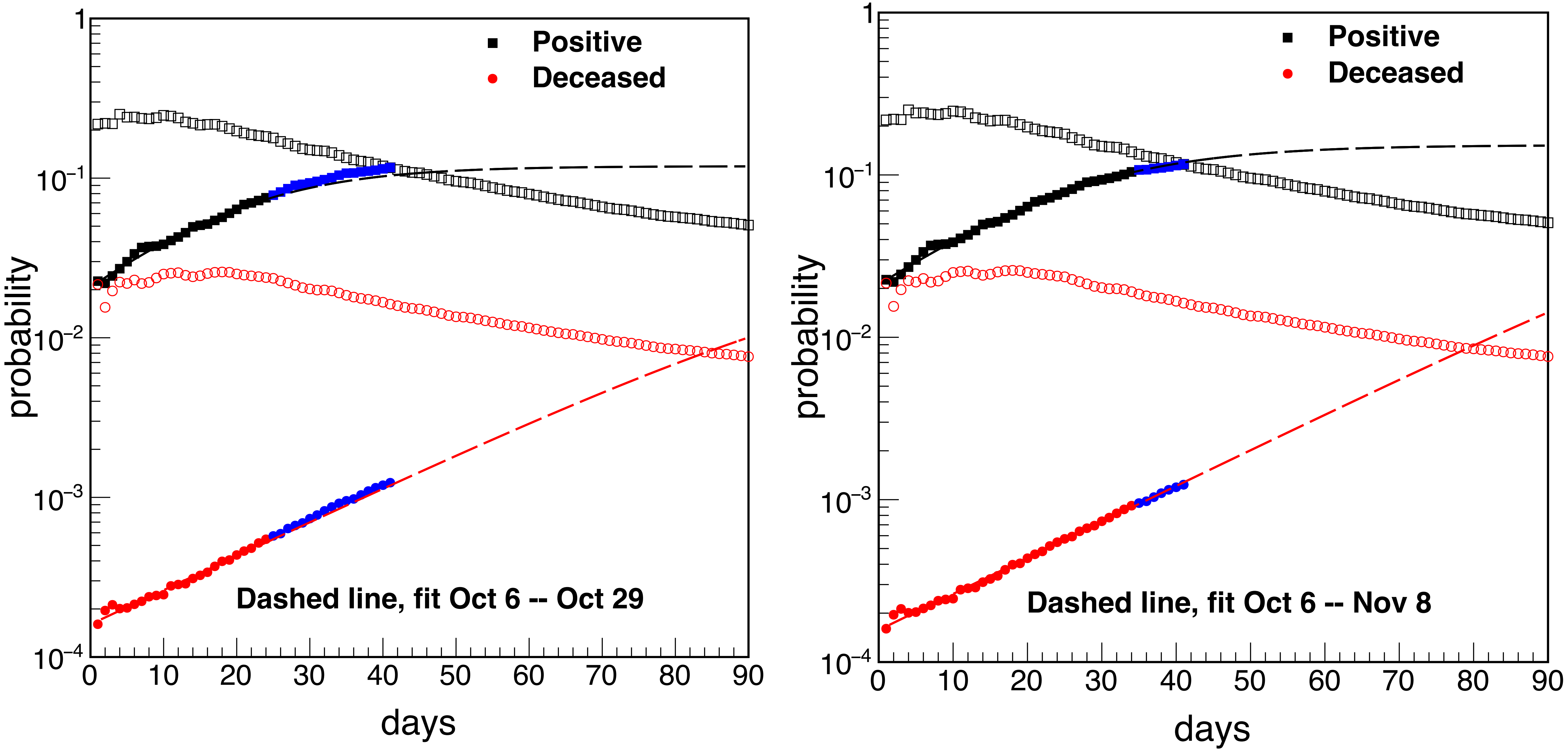


$$\Pi(t) = \frac{d_0 d_\infty}{d_0 + d_\infty e^{-\gamma t}}$$

**open symbols refer to data starting from March 14, 2020 (lockdown)**

#### Two fitting periods:
#### 1) oct6-oct29 (24 days-left panel)
#### 2) oct6-nov8 (34 days-right panel)

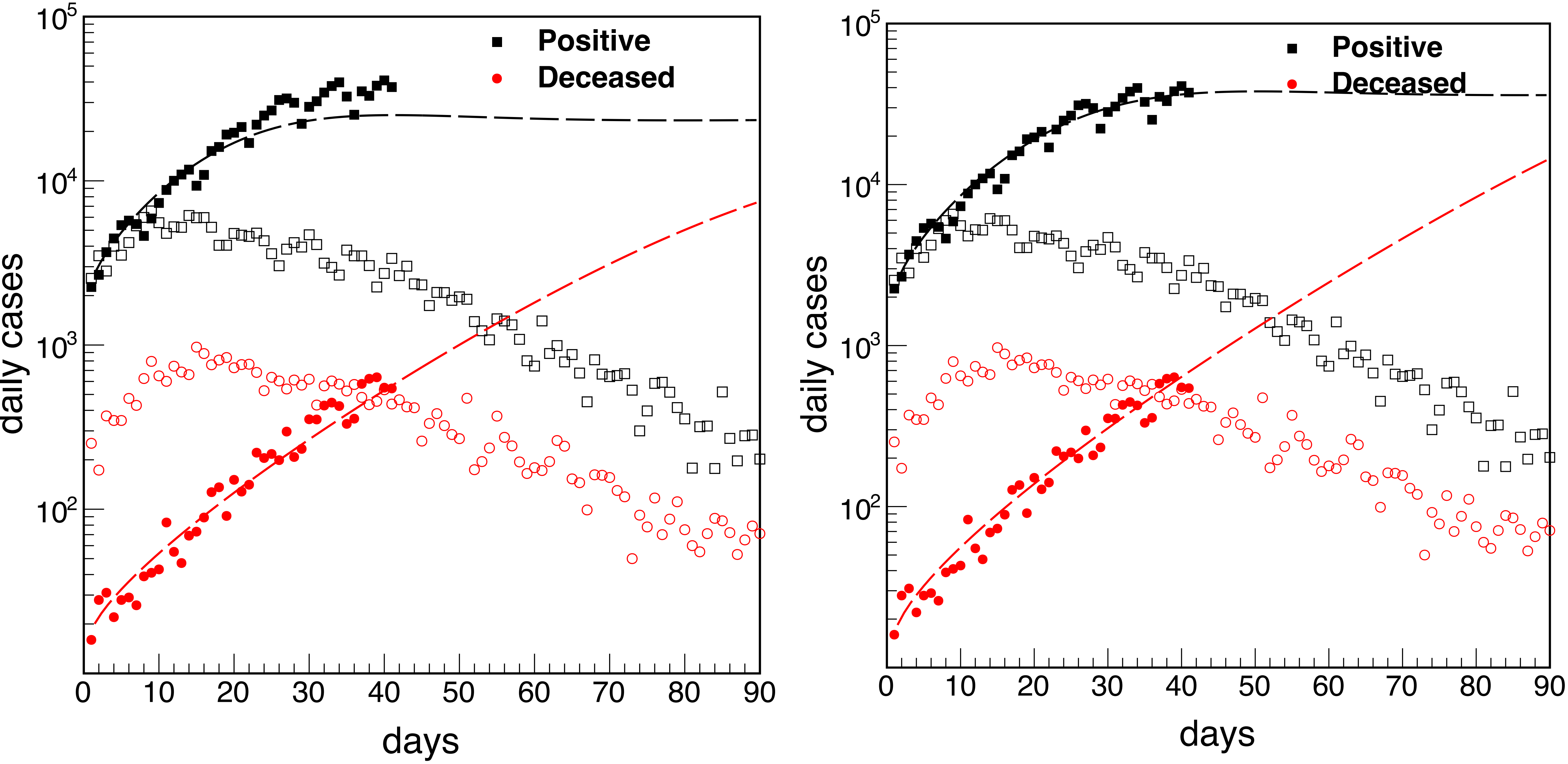


### Time derivative of the #cases

**open symbols refer to data starting from March 14, 2020 (lockdown)**

### Add the study for United Kingdom

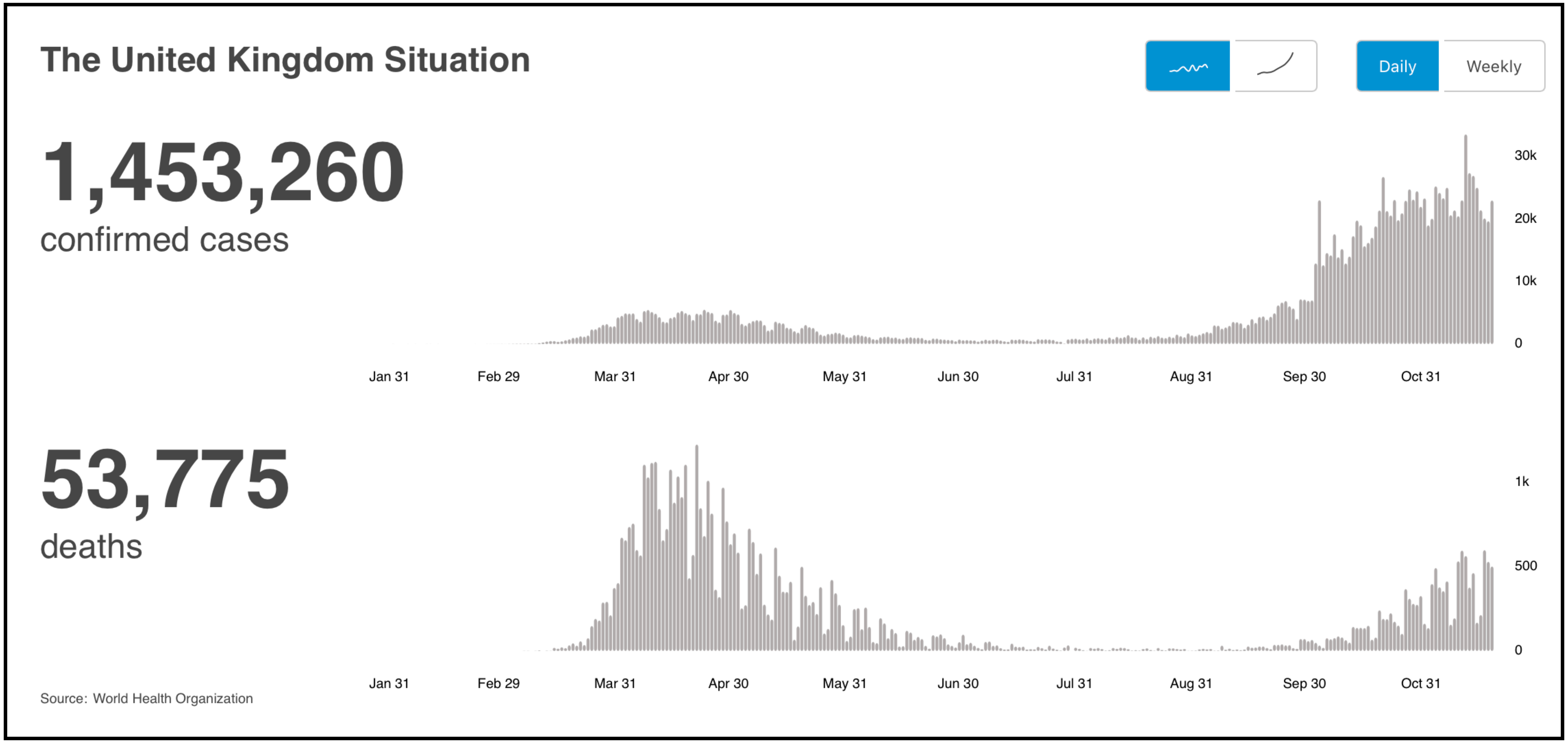


**According to the actual data, we chose Sep 21 as the first day of the second wave of COVID-19 in United Kingdom.**

#### Add the study for United Kingdom

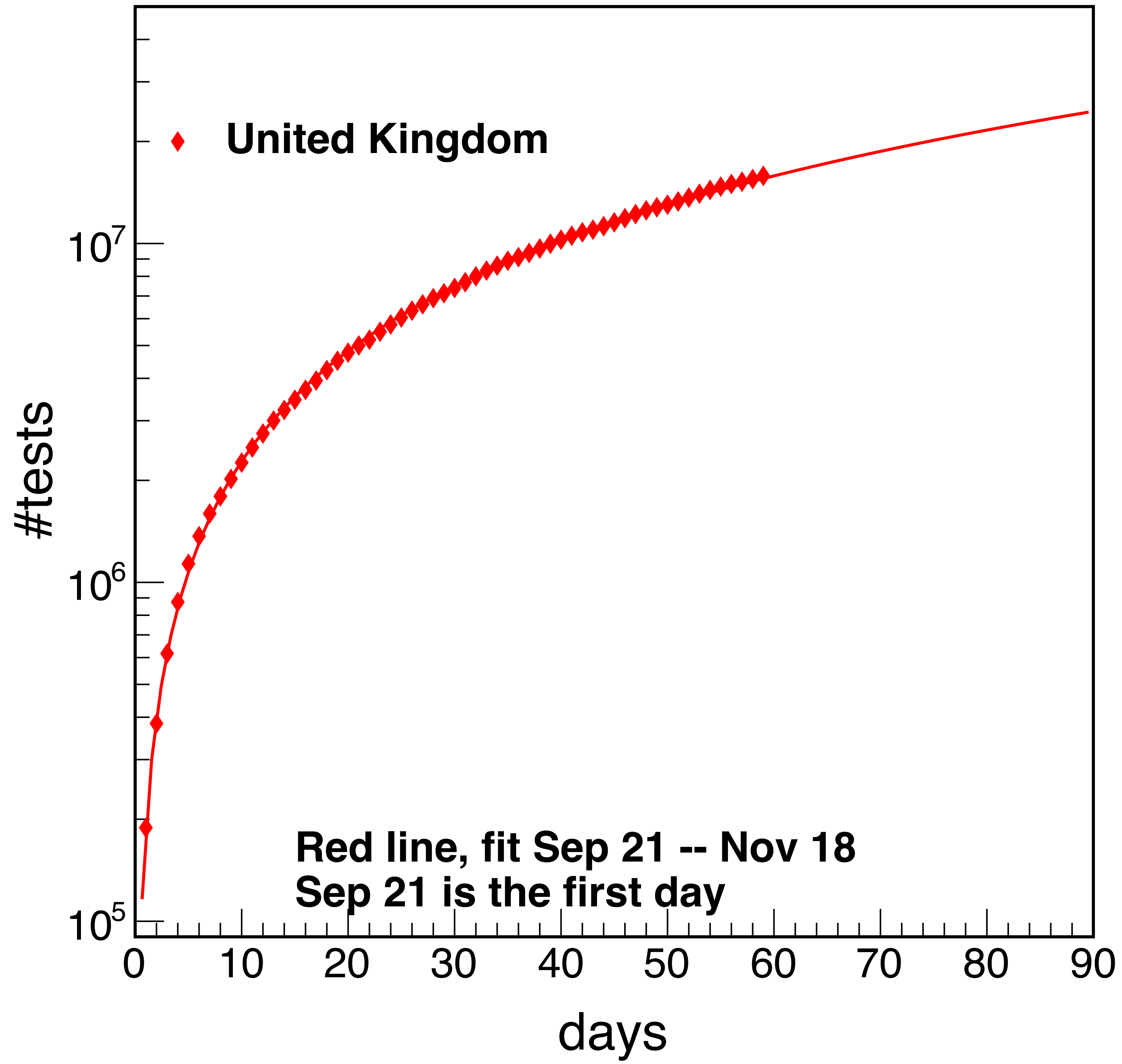


**Fit period**
**Sep 21 - Nov 18**
**Test fit**
**188368,0.00898006,1.0823**

### Add the study for United Kingdom

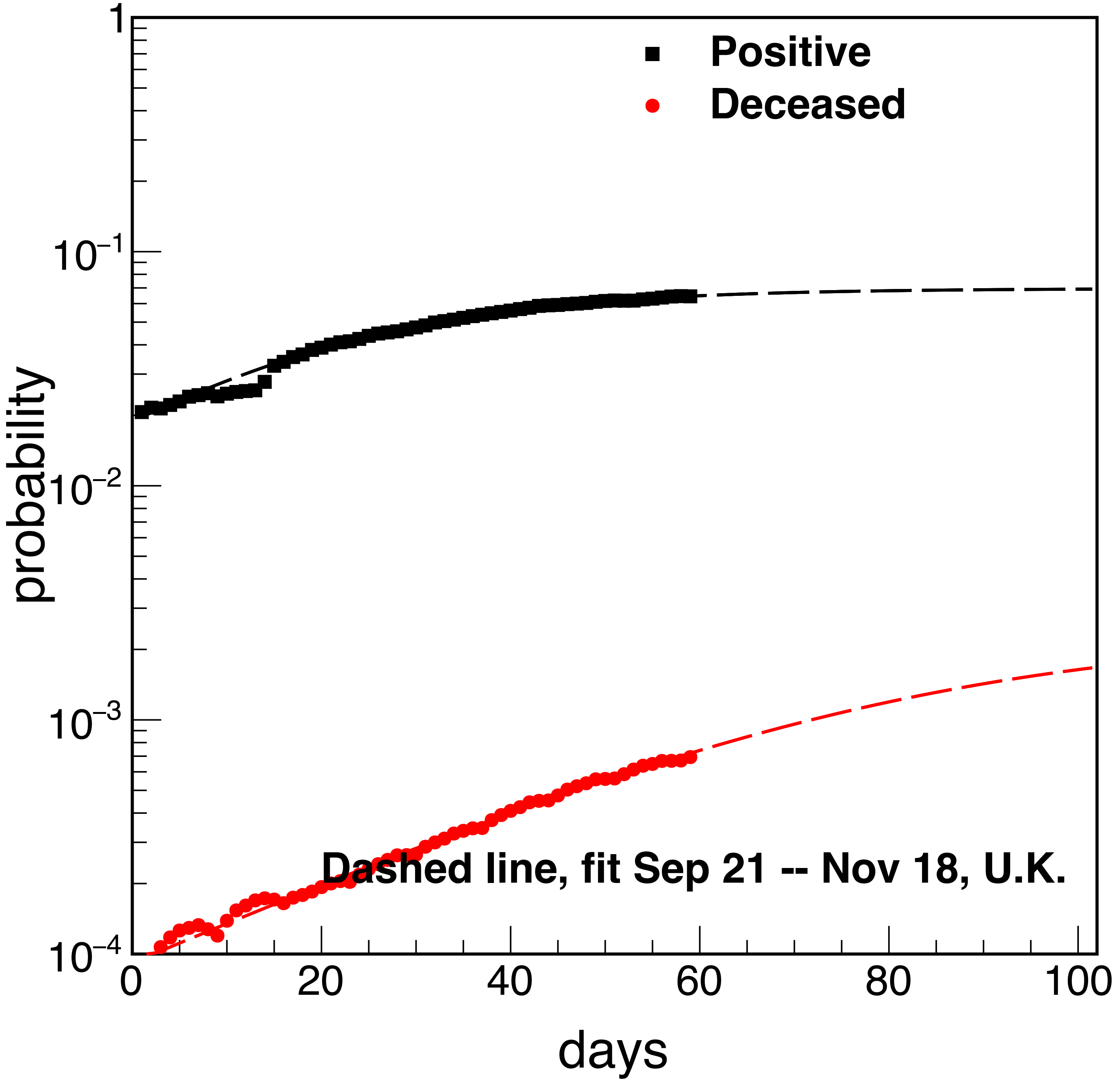


**Fit period**
**Sep 21 - Nov 18**
**Probability fit**
**Pos 0.0258128,0.0696001,0.0600024**
**Dec 9.54838e-05,0.00234158,0.0404792**

### Add the study for United Kingdom, prediction for Dec 31, 2020

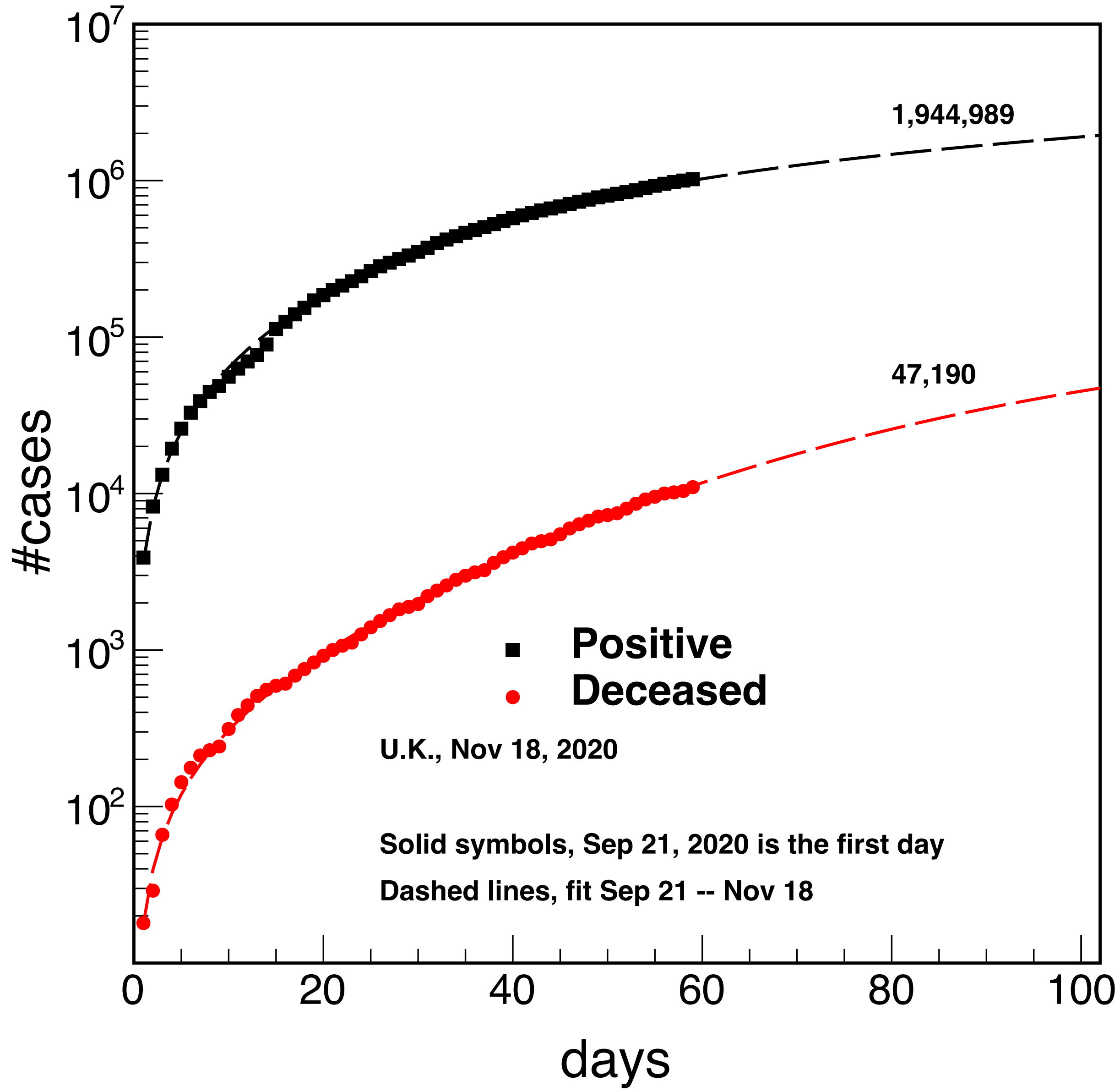


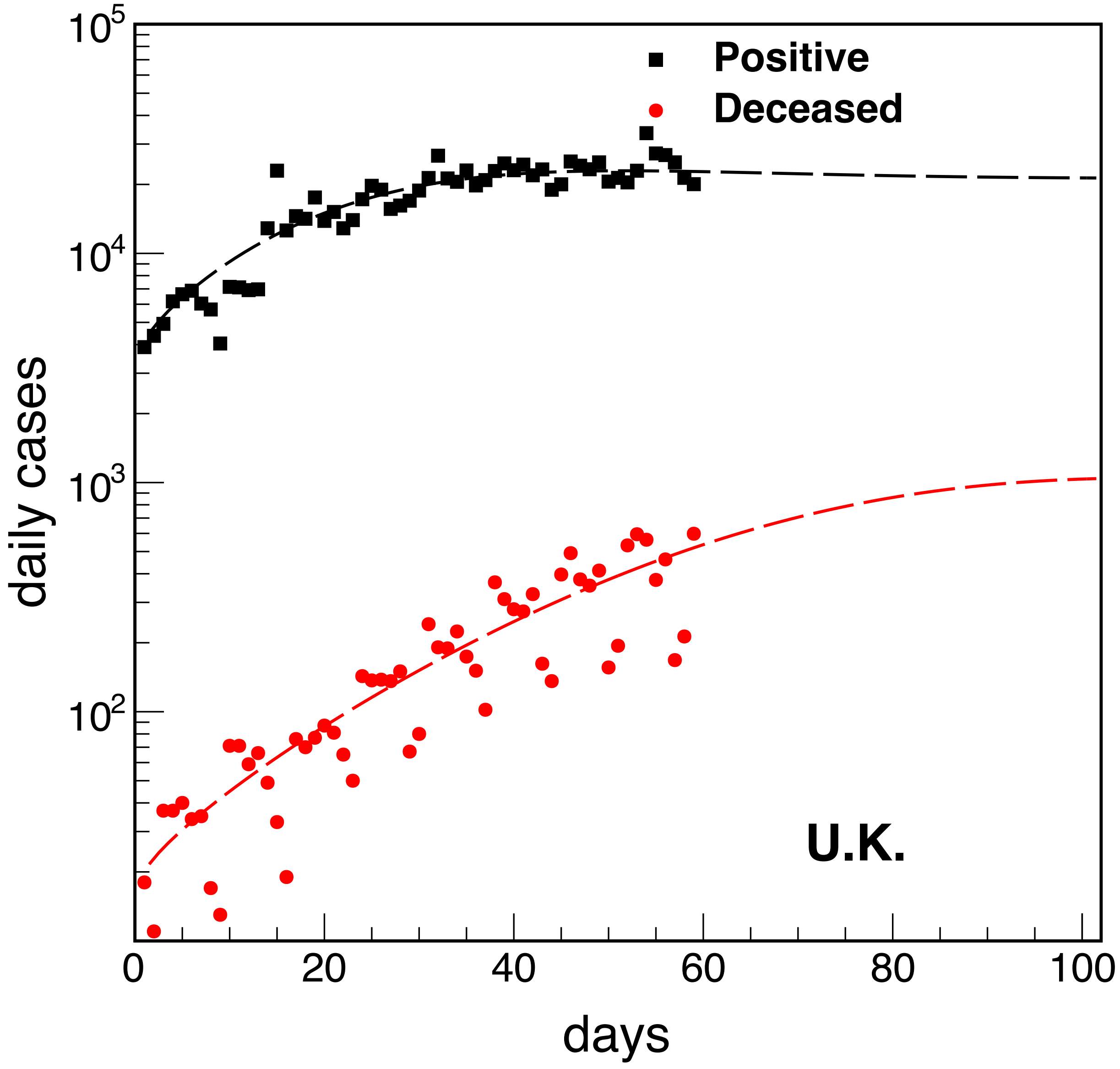
